# Hematological and biochemical predictors of bone marrow metastases in non-hematological malignancies: a clinico-pathological analysis

**DOI:** 10.1101/2020.12.16.20248306

**Authors:** Shruti Agrawal, Rekha Bhandari, Vinay N Gowda, Amit Gupta, Neha Singh, Nilotpal Chowdhury, Shalinee Rao, Sanjeev Kishore, Arvind Kumar Gupta, Harish Chandra

## Abstract

**Background:** Bone marrow metastasis is a significant presentation of many non-hematological malignancies. The present study was conducted with the aim of analysing the clinical, hematological and biochemical parameters of the patients with bone marrow metastases and to determine the parameters which could be possible indicators of bone marrow metastases.

**Material and methods:** Bone marrow aspirate, imprint and biopsy slides of 22 cases with bone marrow metastases were reviewed and the hematological and biochemical data of all these cases was compared with that of 20 controls who had no evidence of metastases.

**Results:** The mean values of mean platelet volume (MPV), neutrophil to lymphocyte ratio (NLR) and serum lactate dehydrogenase (LDH) were found to be significantly different (p<0.001) between the cases and controls. A MPV<8.1 fL, NLR>3.5 and LDH>452 U/L showed high likelihood ratio in predicting bone marrow metastases.

**Conclusion:** Parameters such as MPV, NLR and LDH can be used as efficient and easily available means to predict bone marrow metastasis for an early diagnosis.

## Introduction

Bone marrow examination has traditionally been a routine investigation for various haematological malignancies but marrow involvement by non-hematological malignancies is a relatively uncommon occurrence. The advent of ancilliary techniques such as fluorodeoxyglucose-positron emission tomography (FDG-PET) and bone scans have revolutionized the early and rapid detection of unsuspected cases of bone metastases, however; few cases especially with unknown primary or those presenting with non-specific symptoms are still left undetected and remain a diagnostic challenge for the clinicians. In such cases, bone marrow aspiration (BMA) and trephine biopsy serve to be a sensitive diagnostic tool. Bone marrow biopsy (BMB) also aids in determining the management and prognosis as it is included as a part of staging procedure in most malignancies such as the small round cell tumors.^1,2^ Hence, bone marrow evaluation provides a rapid and sensitive method of detecting metastases, thereby allowing early treatment of the patient and affecting overall survival.

Despite being a sensitive and effective method, bone marrow evaluation may sometimes fail to detect metastases due to a “dry tap” or inadequate BMB. The major objective of this study was to analyze the clinical presentation, as well as hematological and biochemical profile of patients presenting with solid tumor metastases. The study also aimed to identify those hematological and biochemical parameters which could effectively predict bone marrow metastases, thereby prompting the pathologists to search cautiously for abnormal cell clusters in the bone marrow or recommend a repeat procedure if the BMA or BMB are inadequate in such cases.

## Materials and Methods

A retrospective study was conducted in the Department of Pathology to include all the cases of non-hematological malignancies of bone marrow for a period of 28 months from January 2018 to April 2020. Cases of leukemias, lymphomas and plasma cell neoplasms were excluded from the study. The bone marrow aspirate/ biopsy had been done in these cases in view of clinical history of bone pain, radiologically suspected bony lesions, one or more cytopenias not attributable to chemotherapy, or as a part of staging procedure in a known case of malignancy. Twenty cases with a known history of malignancy, but without bone marrow metastases after adequate sampling and radiological screening, were taken as controls. Detailed clinical history of the patients was recorded. Hematological and biochemical values of the patients at the time of bone marrow procedure were noted. Hematological data included haemoglobin levels (Hb), total leukocyte count (TLC), platelet count, absolute neutrophil count (ANC), absolute lymphocyte count (ALC), Mean corpuscular volume (MCV), Red cell Distribution Width (RDW), Mean Platelet Volume (MPV) and Platelet Distribution Width (PDW) which were run on an adequately quality controlled LH750 Beckman Coulter autoanalyser (California, USA). Neutrophil to lymphocyte ratio (NLR) was calculated for every patient. Peripheral blood smear findings were also noted in all cases. Biochemical parameters included serum Alkaline Phosphatase (ALP) and Lactate Dehydrogenase (LDH). Anaemia was defined as Hb value <13 g/dL in males and <12g/dL in females. Leukocytosis was defined as TLC > 11000/uL and leukopenia as TLC <4000/ uL. A platelet count <150×10^3^/ uL was considered as thrombocytopenia. BMA, imprint smears and BMB for all cases and controls were reviewed by two pathologists. Bone marrow procedure was performed under all aseptic precautions from the posterior superior iliac crest in all cases using Salah needle for BMA and Jamshidi needle for BMB. The aspirate and imprint smears were stained with Giemsa stain. The trephine biopsy was stained with Hematoxylin and Eosin after undergoing standard processing techniques comprising of fixation and decalcification. An adequacy criterion of at least 1.5cm length was adopted for all trephine biopsies. Appropriate immunohistochemistry was applied wherever necessary.

All statistical analyses were done using SPSS version 20.0 software. The results were analysed using the independent sample t-test to assess significant difference between the cases and control values with 95% confidence intervals (CI). A p value<0.05 was considered to be statistically significant. Additionally, a receiver operating characteristic curve (ROC) was used to calculate sensitivity, specificity, likelihood ratio and area under curve (AUC) to evaluate the usefulness of a particular variable as a predictive marker of bone marrow metastasis. A cut-off value for each parameter was determined so as to obtain the most optimum values of sensitivity and specificity.

## Results

Of the total 1538 bone marrow procedures done in our institute between January 2018 to April 2020, 22 cases (1.4%) showed bone marrow infiltration by solid tumor. Of these, 12 cases underwent bone marrow procedure as a staging protocol for management of a histologically proven malignancy, 3 were clinically diagnosed as multiple myeloma and remaining underwent a bone marrow examination to evaluate for abnormal blood counts. Bone marrow procedure was done from unilateral site in all the 7 clinically unsuspected patients. Remaining 15 cases underwent bilateral bone marrow examination. A dry tap in BMA was obtained in 6 cases, whereas 8 cases had diluted aspirates. Atypical cells could be seen in the BMA of only 4 cases (18%). However, imprint smears were available for all the cases and revealed the neoplastic cells in 20 cases (90%) (Figure 1). Bone marrow biopsies were adequate in all the cases, ranging in size from 1.5 to 2.2 cm in length. The clinico-hematological and biochemical profile of all the cases with bone marrow metastases are shown in Table 1. The age of the patients with bone marrow metastases ranged from 3 to 84 years with 6 cases belonging to paediatric age group. The demographic profile of the cases and controls are summarised in Table 2. Most common chief complaints of the patients comprised of back pain and difficulty in walking (72%), followed by weight loss and anorexia (64%), and breathlessness and generalised weakness (45%). Other complaints included fever and primary organ specific symptoms such as lower urinary tract symptoms, hematuria and cough. Table 2 shows the histologically confirmed primary malignancy in all the patients, the most common being Ewing’s sarcoma. The histological type of malignancy was determined by the characteristic morphology on BMB and confirmed by appropriate IHC (Figure 1) which was decided after correlating with the clinical complaints and the radiological investigations of the patients. However, in two cases, the primary site could not be established.

**Table 1:**
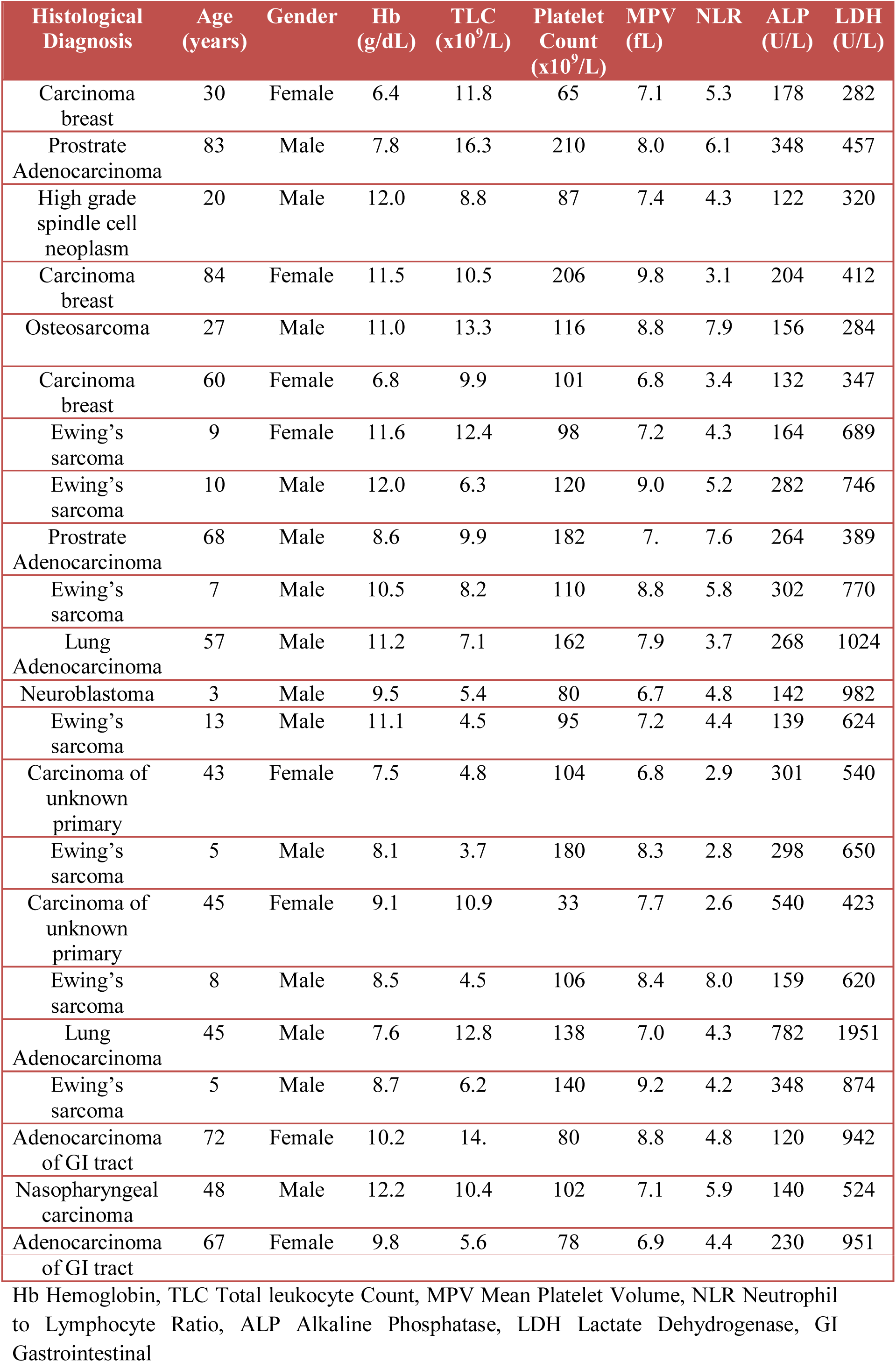
Clinico-hematological and Biochemical profile of the cases with bone marrow metastasis.

**Table 2:** Clinical Features and Primary malignancy of the Patients.

|  | Cases (n=22) | Controls (n=20) |
| --- | --- | --- |
| Gender (Male/Female) | 14/8 | 13/7 |
| Age (Mean±SD) | 36.8±27.8 | 46.7±22.5 |
| Primary site, n (%) |  |  |
| Ewing's sarcoma | 7 (31.8) | 8 (40) |
| Carcinoma, Breast | 3 (13.6) | 2 (10) |
| Adenocarcinoma, Prostate | 2 (9.1) | 2 (10) |
| Adenocarcinoma of GI tract | 2 (9.1) | 1 (5) |
| Adenocarcinoma, Lung | 2 (9.1) | 2 (10) |
| Osteosarcoma | 1 (4.5) | 0 |
| Nasopharyngeal Carcinoma | 1 (4.5) | 0 |
| Neuroblastoma | 1 (4.5) | 0 |
| High Grade Spindle cell Neoplasm | 1 (4.5) | 0 |
| Carcinoma of unknown primary | 2 (9.1) | 0 |
| Carcinoma, Bladder | 0 | 2 (10) |
| Squamous cell carcinoma, Oral cavity | 0 | 2 (10) |
| Adenocarcinoma Gall bladder | 0 | 1 (5) |

SD Standard Deviation, GI Gastrointestinal

**Figure 1.**
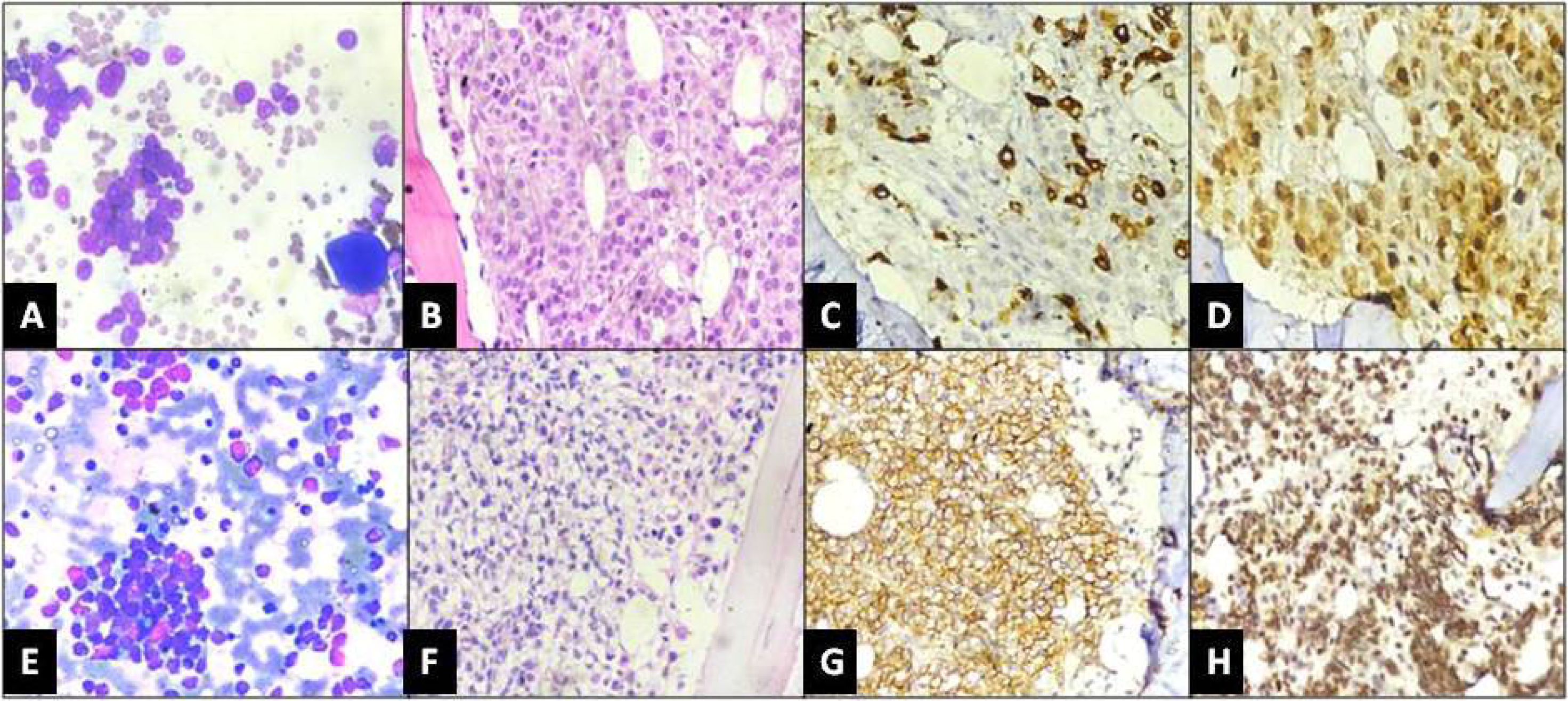
Bone marrow and immunohistochemistry findings in a case of breast adenocarcinoma (A-D) and Ewing’s Sarcoma (E-H). A: Aspirate smear showing clusters of neoplastic cells (x400, Giemsa), B: Trephine biopsy showing sheets and tubules of atypical cells (x400, Hematoxylin & Eosin), C: Tumor cells showing strong cytoplasmic positivity for Mammaglobin and (x400, DAB), D: Gross Cystic Disease Fluid Protein-15 (x400, DAB), E: Imprint smear displaying clusters of small round cells (x400, Giemsa), F: Trephine biopsy showing diffuse infiltration by tumor cells (x400, Hematoxylin & Eosin), G: Tumor cells showing strong membranous positivity for CD99 and (x400, DAB), H: Diffuse nuclear positivity for FLI-1 (x400, DAB)

On analysis of haematological parameters, anaemia was seen in all 22 cases (100%) while leukocytosis was present in only 6 cases (27%). Leukopenia was noted in a single patient only (4.5%). Thrombocytopenia was observed in 16 cases (72%). Peripheral smear examination revealed leukoerythroblastic blood picture in 7 cases (32%), microcytic hypochromic RBCs in 4 cases (18%) and macrocytic anemia and haemolytic blood picture in 2 cases (9%) each. Normocytic normochromic blood picture was observed in remaining 7 cases (32%). The mean values of the different haematological and biochemical parameters in the cases and controls and a comparative difference between the two with respect to bone marrow metastases have been represented in table 3. Significant difference was observed in the values of MPV, ALC, NLR and LDH levels between cases and controls. ROC curve analysis showed that of all the parameters, MPV, NLR and LDH had a significantly higher sensitivity and specificity with a high LR, in determining bone marrow metastasis (Table 4). The ROC curves of these three parameters have been illustrated in figure 2. MPV < 8.1 fL had 63% sensitivity and 90% specificity in cases with bone marrow metastasis as compared to the controls with no evidence of bone marrow metastasis, with a likelihood ratio of 6.3. NLR >3.5 could detect bone marrow metastasis with 77% sensitivity and 85% specificity. Also, an NLR >4.7 has a 100% specificity for predicting bone marrow metastasis. In the biochemical parameters, serum LDH value > 452 U/L carried 68% sensitivity and 90% specificity in diagnosing solid tumor metastasis with a LR value of 6.8.

**Table 3:** Comparison of Hematological and Biochemical parameters between the cases and controls.

| Parameter | Cases (n=22)<br>Mean±SD | Controls (n=20)<br>Mean±SD | P-value | 95% CI |
| --- | --- | --- | --- | --- |
| Haemoglobin (g/dL) | 9.6 ± 1.8 | 10.4 ± 1.3 | 0.11 | -1.8 0.19 |
| TLC ($\times 10^9$/L) | 8.9 ± 3.6 | 7.9 ± 3.0 | 0.32 | -1.1 3.1 |
| Platelet count ($\times 10^9$/L) | 126.9 ± 56.6 | 168.8 ± 96.9 | 0.09 | -90.8 7.1 |
| RBC Count ($\times 10^9$/L) | 3.7 ± 0.87 | 3.8 ± 0.81 | 0.82 | -0.5 0.4 |
| MCV (fL) | 82.3 ± 6.6 | 84.9 ± 6.5 | 0.22 | -6.6 1.5 |
| RDW-CV(%) | 15.4 ± 1.8 | 15.2 ± 1.6 | 0.72 | -0.8 1.2 |
| MPV (fL) | 7.8 ± 0.9 | 9.7 ± 1.1 | <0.001 | -2.6 -1.3 |
| PDW | 9.9 ± 1.9 | 9.6 ± 1.8 | 0.52 | -0.8 1.5 |
| ANC ($\times 10^9$/L) | 6.5 ± 3.0 | 5.1 ± 2.6 | 0.12 | -0.4 3.2 |
| ALC ($\times 10^9$/L) | 2.3 ± 0.7 | 4.7 ± 2.7 | <0.001 | 1.2 3.7 |
| NLR | 4.8 ± 1.6 | 2.3 ± 1.2 | <0.001 | 1.6 3.4 |
| ALP (U/L) | 264.5 ± 162.4 | 186.5 ± 95.2 | 0.07 | -6.2 162.1 |
| LDH (U/L) | 672.7 ± 371.3 | 285.9 ± 123.8 | <0.001 | 210.5 563.1 |

TLC Total Leukocyte Count, RBC Red Blood Cell, MCV Mean Corpuscular Volume, RDW-CV Red cell distribution width-coefficient of variation, MPV Mean Platelet Volume, PDW Platelet Distribution Width, ANC Absolute Neutrophil count, ALC Absolute Lymphocyte Count, NLR Neutrophil to Lymphocyte Ratio, ALP alkaline Phosphatase, LDH lactate dehydrogenase, SD standard deviation, ALP Normal range 30-120 U/L, LDH Normal range 48-247 U/L

**Table 4:** ROC curve Analysis of the Haematological and the Biochemical Parameters.

| Parameter | Cut-off value | Cases (N=22)<br>n(%) | Controls (N=20)<br>n(%) | Sens (%) | Spec (%) | LR | AUC | 95% CI (deLong method) |
| --- | --- | --- | --- | --- | --- | --- | --- | --- |
| Haemoglobin (g/dL) | <9.5 | 12(54) | 4(20) | 50 | 80 | 2.5 | 0.63 | 0.45 to 0.80 |
| TLC (x10<sup>9</sup>/L) | >11.3 | 6(27) | 4(20) | 27 | 80 | 1.4 | 0.58 | 0.40 to 0.76 |
| Platelet count (x10<sup>9</sup>/L) | <101.5 | 14(64) | 10(50) | 68 | 50 | 1.4 | 0.61 | 0.44 to 0.79 |
| RBC Count (x10<sup>9</sup>/L) | <4.0 | 12(54) | 10(50) | 63 | 50 | 1.3 | 0.53 | 0.36 to 0.72 |
| MCV (fL) | <85.1 | 12(54) | 11(55) | 54 | 45 | 0.9 | 0.58 | 0.41 to 0.77 |
| RDW-CV (%) | >14.8 | 11(55) | 10(50) | 50 | 50 | 1.0 | 0.52 | 0.30 to 0.66 |
| MPV (fL) | <8.1 | 14(64) | 2(10) | 63 | 90 | 6.3 | 0.89 | 0.80 to 0.99 |
| PDW | >9.2 | 14(64) | 8(40) | 63 | 60 | 1.6 | 0.55 | 0.36 to 0.73 |
| ANC (x10<sup>9</sup>/L) | >6.1 | 12(54) | 5(25) | 54 | 75 | 2.2 | 0.63 | 0.46 to 0.81 |
| ALC (x10<sup>9</sup>/L) | <2.5 | 13(59) | 5(25) | 77 | 55 | 1.7 | 0.70 | 0.65 to 0.95 |
| NLR | >3.5 | 17(77) | 3(15) | 77 | 85 | 5.1 | 0.90 | 0.80 to 1.0 |
| ALP (U/L) | >205 | 12(54) | 7(35) | 50 | 65 | 1.4 | 0.68 | 0.52 to 0.85 |
| LDH (U/L) | >452 | 15(68) | 2(10) | 68 | 90 | 6.8 | 0.90 | 0.81 to 0.99 |

ROC Receiver Operating Characteristic curve, TLC Total Leukocyte Count, RBC Red Blood Cell, MCV Mean Corpuscular Volume, RDW-CV Red cell distribution width-coefficient of variation, MPV Mean Platelet Volume, PDW Platelet Distribution Width, ANC Absolute Neutrophil count, ALC Absolute Lymphocyte Count, NLR Neutrophil to Lymphocyte Ratio, ALP alkaline Phosphatase, LDH lactate dehydrogenase, Sens Sensitivity, Spec Specificity,
LR likelihood ratio, AUC Area under ROC curve, CI Confidence Interval, ALP Normal range 30-120 U/L, LDH Normal range 48-247 U/L

**Figure 2:**
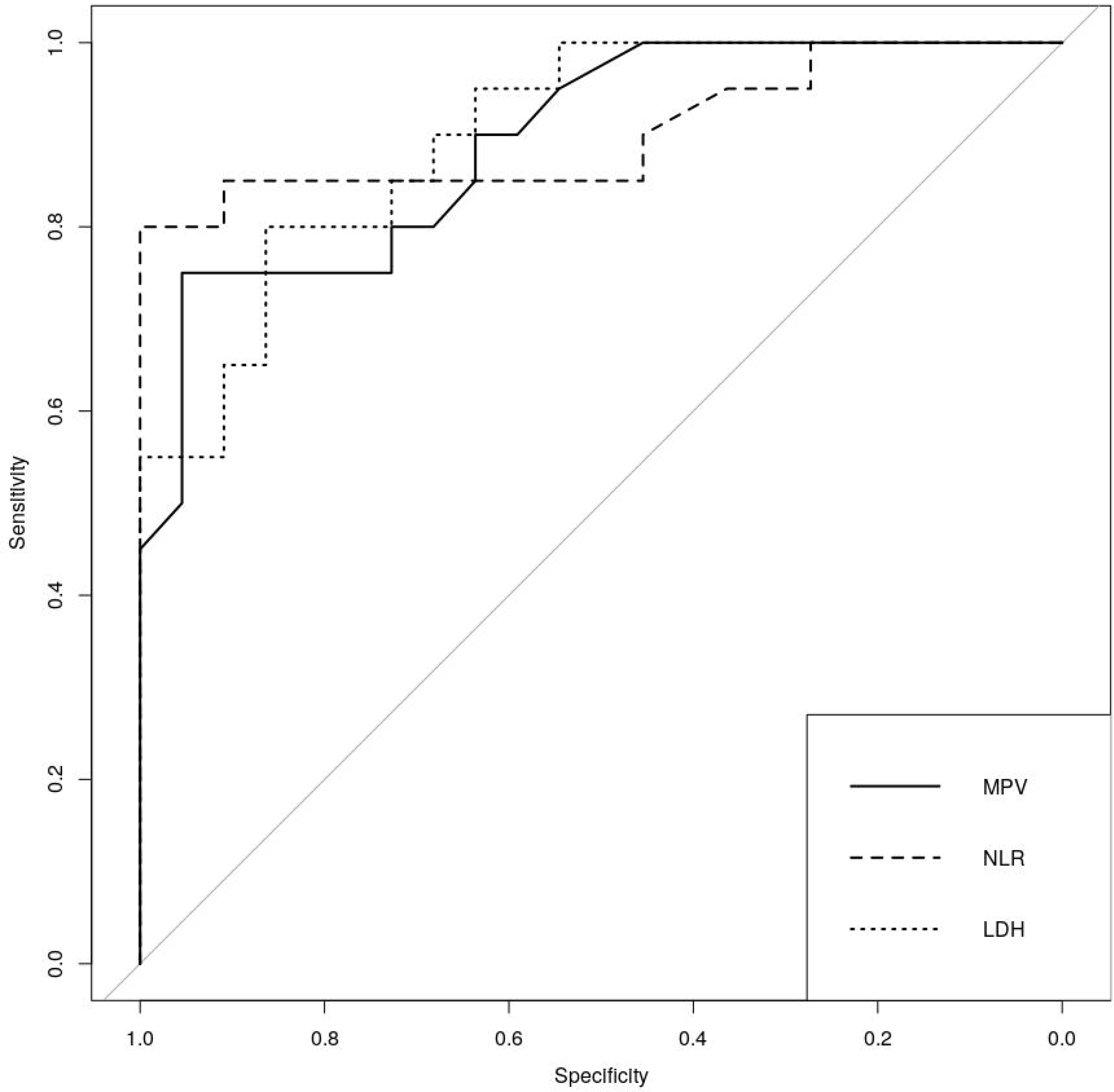
Plot showing Receiver Operating Characteristic curves for mean platelet volume (MPV), neutrophil to lymphocyte ratio (NLR) and lactate dehydrogenase (LDH). All the three curves show high area under the curve.

## Discussion

Bone marrow evaluation is the most useful and sensitive technique for detecting bone marrow metastases in non-hematological malignancies. Infiltration of bone marrow by solid tumors is an important presentation. In our study, only 1.4% of all the bone marrow procedures revealed solid tumor metastases over a period of 28 months. Evaluation of BMA and trephine biopsy specimens are being utilised increasingly over the past few decades to determine the accurate staging of certain malignancies.^1,2^ In the present study, bone marrow involvement aided the clinicians in upstaging the tumor and planning the management accordingly, in 12 cases (54%). However, the major diagnostic utility of the bone marrow examination was evident in those cases where a primary malignancy was completely unsuspected clinically (32%) or the patient was being worked up for an initial diagnosis of multiple myeloma (14%), thereby providing valuable information to the clinicians. Importance of a meticulous bone marrow evaluation has also been reiterated in literature, especially in cases where bone marrow metastases is the initial presentation of solid tumors.^1-4^ Although bone marrow examination is quite sensitive in detection of metastases, many a times, the adequacy of the material obtained proves to be a major limiting. A “dry tap” in BMA, where no material is aspirated despite multiple attempts or a scanty non-diagnostic yield is obtained, may result from extensive bone marrow fibrosis or hypercellularity in a metastatic involvement of bone marrow. In such cases, examination of the imprint smears proves beneficial in detecting the atypical cells which might otherwise be missed out.^5,6^ In the present study, tumor cells could be picked up in only 18% of the BMA smears whereas 14 aspirates were either diluted or yielded a dry tap. However imprint smears helped in identifying the atypical cells in 90% of the cases. Gupta et al reported the diagnostic utility of BMA in 76% of BMA and 97% of imprints in non-hematological disorders.^6^ Therefore a careful and diligent preparation of imprint/ touch smears from trephine biopsies is necessary for accurately replicating core biopsy findings and increasing the sensitivity of diagnosing metastases.

Similarly, for BMB, the length of the biopsy is a crucial factor was assessing the adequacy. WHO (2016) recommends a minimum length of 1.5cm for BMB to be adequate enough for evaluating any neoplasm.^7^ The size criteria ensures precise detection of even those lesions which tend to involve the bone marrow focally, especially metastatic tumors. In our study, the trephine biopsies were adequate and revealed the atypical cells in 100% cases. This is consistent with the findings in other studies which have reported the diagnostic superiority of BMB over BMA in cases of solid tumor metastases.^5,6,8^ Additionally, bilateral bone marrow examination has been advocated to increase the diagnosis of marrow involvement in many haematological and non-hematological malignancies and was done in 15 cases (68%) in our study.^9^

Clinically, the most frequent complaint of the patients in our study was back pain associated with disabled movements in 72% of the patients. This finding is in accordance to previous studies conducted by Chandra et al and Wong et al.^10,11^ Although bone marrow metastases is known to occur in many solid tumors, the most common primary tumors that metastasize to bone marrow are lungs, breast, stomach and prostate.^12-14^ Overall most frequent primary malignancy in the present study was Ewing’s sarcoma (31.8%) followed by breast carcinoma (13.6%). There were 2 cases (9.1%) each of the prostate, gastrointestinal and lung adenocarcinoma. Anner et al however reported metastasis from neuroblastoma (48.2%) as the most frequent occurrence followed by Ewing’s sarcoma (35.5%).^14^ Gupta et al observed gastric adenocarcinoma to be the most common primary tumor.^6^ In the present study, metastatic adenocarcinomas from various primary sites were evident on histology of trephine biopsy in 9 cases (40.9%). However, despite an extensive radiological and immunohistochemical analysis, the primary site could not be established in two cases.

The haematological parameters in cases of bone marrow metastases usually reveal one or more cytopenias.^1,3,4^ In the present study, anemia and thrombocytopenia were documented in 100% and 72% of the cases, with the leukocyte count being within normal range in most patients (mean 8.9×10^9^/L). This finding is similar to the observations of Ozkalemkas et al, who reported anemia and thrombocytopenia in all of their cases. In fact, one of the main reasons for bone marrow evaluation in their study was unexplained cytopenias in cases of unsuspected non-hematological malignancies. They also concluded that presence of leukoerythroblastic blood picture and microangiopathic haemolytic anemia should trigger a necessity for bone marrow evaluation.^1^ In the present study, leukoerythroblastic blood picture was observed in 32% cases while haemolytic anemia was recorded in 9% cases only. Delsol et al reported leukoerythroblastosis in 44% of the patients with bone marrow metastases and considered this haematological finding as a consistent feature of solid tumor metastases. ^15^

An important objective of the present study was to determine the haematological and biochemical parameters which could possibly predict the event of bone marrow metastases in cases of non-hematological malignancies. At times, the neoplastic infiltration in the bone marrow may be quite indistinct such that the cells may not be distinguished on histomorphology alone. In these cases, use of appropriate IHC markers or even molecular studies facilitates adequate visualisation of the malignant cells.^9,16^ However, in few cases, even after the application of ancillary techniques, bone marrow metastases may not be detected, especially when the involved lesion is very focal. In such instances, a knowledge of haematological or biochemical predictive variables could give a clue to the pathologist to consider a careful second examination of the bone marrow. In the present study, a statistically significant difference was observed between the cases with bone marrow metastasis and the controls with uninvolved marrow with respect to MPV, ALC, NLR and LDH. On ROC curve analysis, an MPV value <8.1 fL had a specificity of 90% and a high likelihood ratio in predicting bone marrow metastasis. The exact reason for decrease in platelet volume in bone marrow metastases is unknown, yet it has been proposed that thrombopoietin production by marrow stromal cells may be affected due to interaction with metastatic cells, which results in deranged maturation of the megakaryocytes and hence influences the platelet volume.^17^ As documented by Aksoy et al, MPV <7.4 fL had a sensitivity of 82.7% and a specificity of 89.6% for bone marrow metastasis.^18^ Though mean values of ALC were significantly different (p<0.001) between the cases and the controls, the specificity and LR derived from the ROC curve analysis were not significant. However, NLR >3.5 showed a moderate sensitivity (77%) and a good specificity (85%), with a high positive LR in predicting bone marrow metastasis. Higher neutrophil counts and lower lymphocyte counts in the cases with bone marrow metastasis as compared to the controls resulted in higher NLR. A relation between systemic inflammation and tumorigenesis is well known. Tumor cells stimulate release of granulocyte-colony stimulating factor which triggers neutrophila, thereby promoting tumor growth and angiogenesis through production of various cytokines. On the other hand, lymphocytes play an important role in providing anti-tumor immunity.^19-21^ Caliskan et al recorded a higher NLR in patients with bone metastases as compared to those without bone metastases.^20^ Similarly, Wang et al reported that a high NLR (>3) was independently associated with worse prognosis in patients with malignant bone metastases.^21^

Serum LDH >452 U/L showed a 90% specificity and LR of 6.8 for bone marrow metastases in our study. This was in concordance with other studies which also demonstrated the positive correlation of serum LDH values with marrow involvement by metastases.^1,22,23^ Hamrick et al concluded that patients with normal serum LDH along with normal bone scans had <5% chance of marrow involvement.^22^ Other variables such as anemia, thrombocytopenia, elevated RDW, PDW and ALP have also been shown to indicate bone marrow metastases in various studies, however no such correlation was seen in our study.^1,11,23^ The limitation of this study has been a small number of cases and inclusion of a spectrum of malignancies. Also, this was an exploratory study, with the controls being taken by convenience sampling. This study needs to be validated by a larger study to confirm the diagnostic accuracy of MPV, NLR and LDH as well as study the correlation of tumor type and the prognosis of the patient with the various haematological and biochemical parameters.

## Conclusion

Metastatic involvement of bone marrow by solid tumors is a crucial finding and mandates a careful bone marrow examination, aided by a judicious use of immunohistochemistry panel in arriving at a diagnosis. Parameters such as mean platelet volume, neutrophil to lymphocyte ratio and serum lactate dehydrogenase not only serve as rapid and easily available methods to predict the occurrence of bone marrow metastases, but also warrant a meticulous search for the presence of neoplastic cells, to avoid a false diagnosis.

## Data Availability

All the data mentioned in the manuscript including details of all the cases and controls involved in the study have been mentioned in the manuscript itself.

## Description of contributions

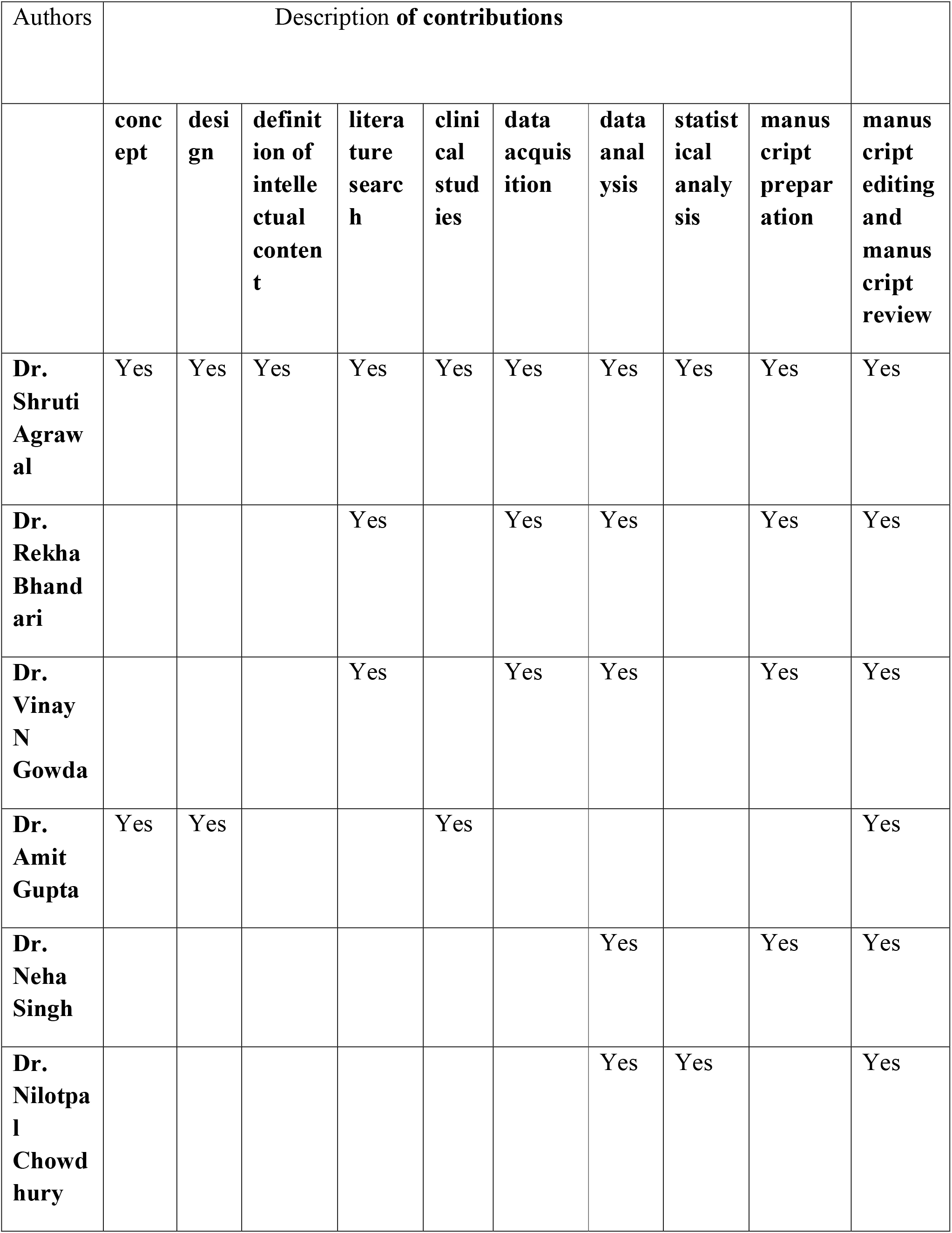
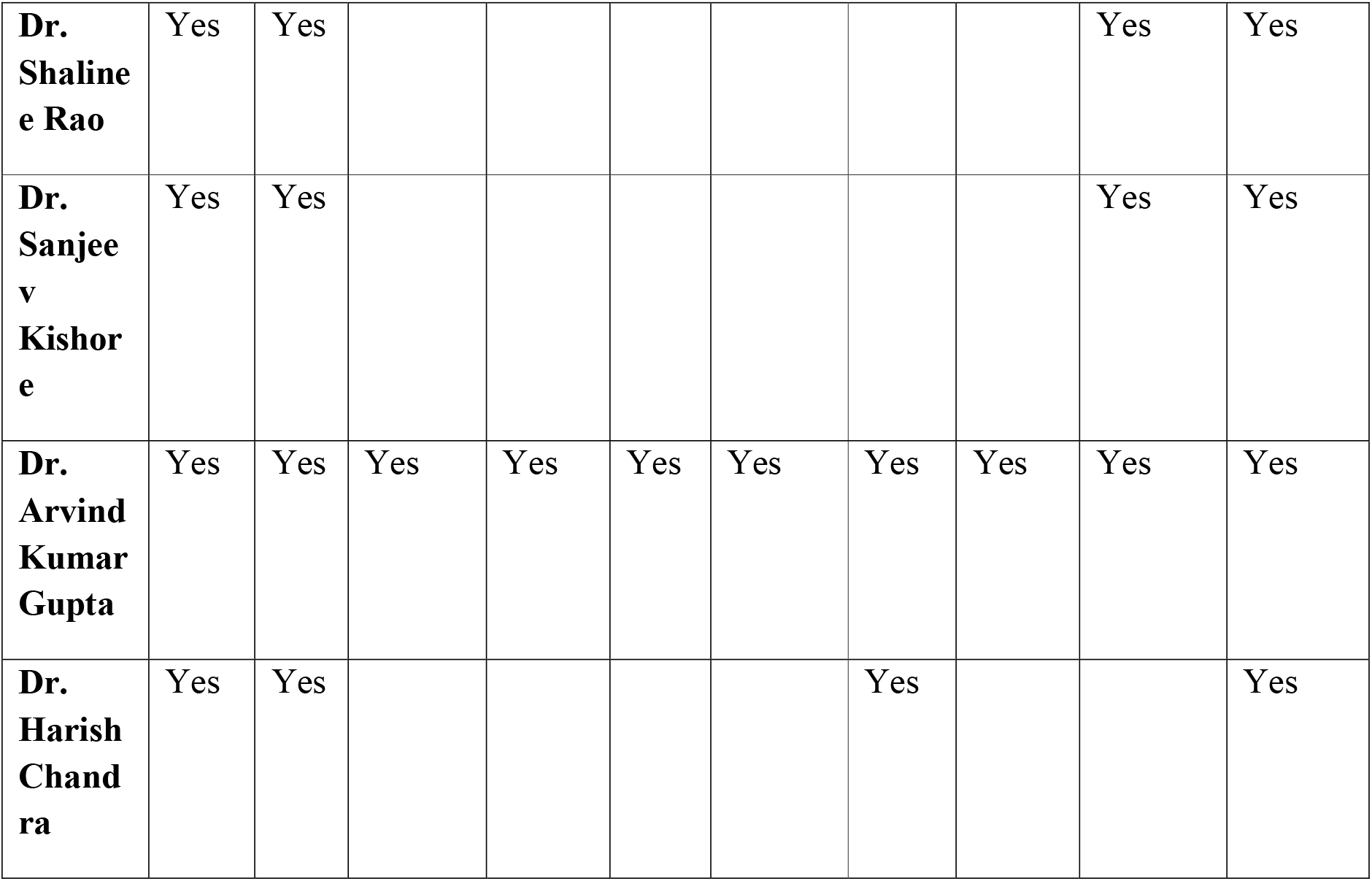

| Authors | Description of contributions |  |  |  |  |  |  |  |  |  |
| --- | --- | --- | --- | --- | --- | --- | --- | --- | --- | --- |
|  | concept | design | definition of intellectual content | literature search | clinical studies | data acquisition | data analysis | statistical analysis | manuscript preparation | manuscript editing and manuscript review |
| **Dr. Shruti Agrawal** | Yes | Yes | Yes | Yes | Yes | Yes | Yes | Yes | Yes | Yes |
| **Dr. Rekha Bhandari** |  |  |  | Yes |  | Yes | Yes |  | Yes | Yes |
| **Dr. Vinay N Gowda** |  |  |  | Yes |  | Yes | Yes |  | Yes | Yes |
| **Dr. Amit Gupta** | Yes | Yes |  |  | Yes |  |  |  |  | Yes |
| **Dr. Neha Singh** |  |  |  |  |  |  | Yes |  | Yes | Yes |
| **Dr. Nilotpal Chowdhury** |  |  |  |  |  |  | Yes | Yes |  | Yes |
| **Dr. Shaline Rao** | Yes | Yes |  |  |  |  |  |  | Yes | Yes |
| **Dr. Sanjeev Kishore** | Yes | Yes |  |  |  |  |  |  | Yes | Yes |
| **Dr. Arvind Kumar Gupta** | Yes | Yes | Yes | Yes | Yes | Yes | Yes | Yes | Yes | Yes |
| **Dr. Harish Chandra** | Yes | Yes |  |  |  |  | Yes |  |  | Yes |

